# The association between malnutrition and cognitive development in infancy as manifest in EEG functional connectivity and power spectral density

**DOI:** 10.1101/2024.09.11.24313505

**Authors:** Berit Hartjen, Shahria Kakon, Navin Rahman, Garrett Greaves, Wanze Xie, Fahmida Tofail, Rashidul Haque, Charles A. Nelson

## Abstract

Malnutrition, particularly undernutrition, is a critical global health challenge, contributing to nearly half of all deaths among children under 5 and severely impacting physical and mental health, along with neural and cognitive development. Prior research by Xie et al. (2019a) linked growth faltering to altered EEG functional connectivity (FC) at 36 months and poorer cognitive outcomes at 48 months; however, no associations were found at 6 months for EEG measures or at 27 months for cognitive outcomes. Our study investigates these relationships in a sample of 12-month-old infants in Dhaka, Bangladesh, using various growth measurements (height/length-for-age, weight-for-age, weight-for-height/length, head-circumference-for-age, and mid-upper-arm-circumference-for-age z-scores) as indicators of nutritional status. Brain development was assessed through EEG, focusing on power spectral density (PSD) and FC, while cognitive development was evaluated with the Bayley Scales of Infant and Toddler Development, Fourth Edition. Our findings reveal that, at 12 months, growth faltering, indicative of undernutrition, was associated with reduced PSD, while initial correlations with increased FC did not remain significant after false discovery rate (FDR) correction. PSD was further positively linked to cognitive development, but associations with FC were not significant post-correction. Notably, EEG PSD in the theta and alpha bands mediated the relationship between malnutrition and behavioral outcomes. These results underscore the early impact of malnutrition on brain development, highlighting the importance of PSD in understanding neural development in this context. Our study emphasizes the need for early intervention and continuous monitoring to mitigate the adverse effects of malnutrition on infant brain and cognitive development.

**RESEARCH HIGHLIGHTS:**

- EEG power spectral density (PSD) in the theta and alpha frequency bands at 12 months sheds light on undernutrition’s impact on cognitive development.
- Better nutritional health, indicated by higher height/length-for-age (HAZ), leads to increased theta and alpha PSD, contributing to better cognitive outcomes.
- Initial correlations between undernutrition, increased EEG functional connectivity (FC), and poorer cognitive outcomes did not remain significant after FDR correction.
- Findings highlight the critical role of nutritional interventions in early childhood to support optimal neural and cognitive development.

## 1 INTRODUCTION

Nutrition profoundly influences neural and cognitive development throughout the lifespan, particularly during critical periods in the first 1000 days of life. According to the World Bank (2024), approximately 250 million children under the age of five in low- and middle-income countries do not reach their developmental potential due to poverty and malnutrition. Malnutrition, defined as deficiencies, excesses, or imbalances in energy and/or nutrient intake, is broadly classified into two categories: (1) undernutrition, comprising stunting (low height/length-for-age – HAZ), wasting (low weight-for-height/length – WHZ), underweight (low weight-for-age – WAZ), and micronutrient-related malnutrition; and (2) overweight, obesity, and diet-related noncommunicable diseases (WHO, 2024a). Malnutrition, especially undernutrition, poses remarkable risks to the developing nervous system, potentially causing permanent impairments in brain functions. While genetic factors play a key role in brain development, environmental influences, such as nutrition, are equally critical (Georgieff et al., 2018).

In 2022, Asia reported the highest prevalence of malnutrition, with over half of all stunted children, 70% of wasted children, and 48% of overweight children under five residing in this region (WHO, 2023). In Bangladesh, a densely populated lower middle-income country in South Asia, 24% of all children under five are stunted, 22% are underweight, and 11% are wasted (NIPORT & ICF, 2023). This underscores the urgent need for targeted nutritional interventions in this vulnerable population. Despite the recognized importance of adequate nutrition for brain and cognitive development, the specific mechanisms linking early childhood malnutrition to behavioral outcomes remain insufficiently explored, particularly in low- and middle-income countries where malnutrition is most prevalent. This study aims to investigate the associations between various growth metrics – used as proxies for undernutrition – and cognitive development in Bangladeshi children, focusing on how brain activity and functional connectivity mediate these relationships.

The human brain is a complex organ with distinct regions and processes, each with specific nutrient requirements throughout development (Georgieff et al., 2018). During early development, the brain evolves from a state of high plasticity and functional non-specificity to greater specialization but reduced malleability. The first 1,000 days of life represent a critical period of rapid brain growth, where the timely provision of essential nutrients is vital for optimal neurodevelopment (Georgieff et al., 2018). Key brain structures begin to form in the first weeks of gestation, followed by neural network formation and myelination, processes dependent on specific nutrients at critical stages. Nutritional deficiencies can lead to enduring impairments, including structural deficits, persistent neurochemical and electrophysiological abnormalities, and altered gene expression, potentially resulting in long-term cognitive deficits, increased disease susceptibility, and heightened risk of neuropsychiatric disorders (Mattei & Pietrobelli, 2019). Malnutrition also contributes to broader social and economic disparities. In Bangladesh, malnourished children are 26% less likely to enroll in school on time and 31% less likely to attend an age-appropriate grade compared to their well-nourished peers (Khanam et al., 2011).

Evidence shows that malnutrition can directly impair brain development. A post-mortem histological study by Cordero et al. (1993) demonstrated that, in the motor cortex of undernourished infants, there is reduced arborization and span of basilar dendrites, suggesting compromised synaptic connections and cortical development. Recent MRI studies have reported cerebral atrophy and dilated ventricles as common structural abnormalities in the brains of malnourished infants (Ayaz et al., 2023). Conversely, greater brain volume has been linked to higher intake of breast milk, calories, and lipids, underscoring nutrition’s role in brain development. A study on nutrient-myelin interaction identified a developmental window between 6 to 20 months where nutrient intake most strongly correlates with brain maturation (Schneider et al., 2023). Collectively, these findings confirm the detrimental effects of malnutrition on brain development, especially in early life.

Low-cost neuroimaging techniques such as electroencephalography (EEG) are particularly valuable in studying malnutrition’s effects on brain development. Other neuroimaging methods, like functional magnetic resonance imaging (fMRI) and functional near-infrared spectroscopy (fNIRS), face cost and accessibility barriers in low-resource settings where malnutrition is prevalent (Galler et al., 2021). EEG provides a direct measure of neural activity by capturing synaptic currents, offering a more immediate and temporally precise understanding of brain function than hemodynamic-based methods like fMRI and fNIRS. The Barbados Nutrition Study of the 1970s collected extensive EEG data from children exposed to protein-energy malnutrition (PEM) during their first year of life. Recent analyses of this data reveal that PEM is associated with significant decreases in alpha activity and insufficient decreases in beta activity in the brain, suggesting delayed brain development (Bringas-Vega et al., 2019; Bosch-Bayard et al., 2022). Additionally, findings show that early malnutrition has enduring negative effects on brain function, persisting for up to 50 years (Razzaq et al., 2022). This research highlights the potential of EEG biomarkers to identify children at risk of neurodevelopmental impairments due to malnutrition, facilitating early intervention in low-resource settings during critical and sensitive period of brain development.

While the Barbados Nutrition Study provided valuable insights into the effects of malnutrition on the developing brain, as well as long-term neurocognitive effects, it did not fully explore the early cognitive consequences, or the underlying neural mechanisms involved. The Bangladesh Early Adversity Neuroimaging (BEAN) project, conducted in the same urban slum of Dhaka as this study, found that brain function might mediate the link between malnutrition and cognitive outcomes. Specifically, in an older cohort, lower HAZ was associated with increased FC (hyperconnectivity) in the theta and beta frequency bands at 36 months, which was further linked to poorer cognitive outcomes at 48 months. Interestingly, however, no such associations between HAZ, FC at six months, and cognitive outcomes at 27 months were observed in a younger cohort. Longitudinal path analysis in the older cohort suggested that FC mediates the effect of HAZ on cognition, implying that brain FC could be a neural pathway through which malnutrition impacts cognitive development. These findings also suggest that elevated FC in stunted children may result from delayed synaptic pruning, compensatory neural mechanisms, or immature brain network organization (Xie et al., 2019). In another study by the BEAN project, growth faltering – assessed by stunting, underweight and wasting as indicators malnutrition – was found to predict brain morphometry in six-year-old children, particularly in white matter and subcortical volumes, mediating the relationship between diminished growth and lower full-scale IQ (Turesky et al., 2020).

As these studies illustrate, malnutrition has significant implications not only for brain development but also for cognitive outcomes over both the short and long term. Although fewer studies examine both brain and cognitive development in this context, evidence consistently shows that malnutrition adversely affects cognitive development. For example, in a large cohort of Nepalese children, low birth weight was associated with poorer performance on intelligence tests, motor skills, and executive function measures at seven to nine months (Christian et al., 2014). Similarly, a study in Thailand found that infant length at birth and during the first year was positively correlated with IQ scores at nine years (Pongcharoen et al., 2012). In India, research among children with severe acute malnutrition revealed that 75% exhibited motor delays, 75% showed language delays, 63% had cognitive delays, and 63% experienced overall developmental delays, with an average lag of four to seven months in total developmental age (Khandelwal et al., 2020). However, some studies suggest that cognitive and neurodevelopmental deficits resulting from undernutrition may be reversible if nutritional status improves before age eight (Suryawan et al., 2022).

Despite the extensive literature demonstrating malnutrition’s adverse effects on brain and cognitive development, there remains a gap in understanding the mechanisms underlying these relationships. Most studies either examine malnutrition’s effects on brain development or cognitive outcomes but rarely integrate these findings to provide a comprehensive view. This study aims to address this gap by combining low-cost neuroimaging techniques with comprehensive behavioral assessments. While prior research has shown that brain FC may mediate malnutrition’s impact on cognition by 36 months, little evidence exists on when these relationships emerge. Our study investigates these associations as early as 12 months, using various growth measures (HAZ, WAZ, WHZ, as well as head-circumference-for-age and mid-upper-arm-circumference-for-age z-scores) as proxies for nutritional status. Brain development was assessed using EEG, focusing on both FC and power spectral density (PSD) across multiple frequency bands. This dual approach enables a more nuanced understanding of malnutrition’s impact on different aspects of brain development. Cognitive development was evaluated using the Bayley Scales of Infant and Toddler Development, Fourth Edition. Mediation analyses were conducted to test whether brain activity and/or connectivity mediated the relationship between nutritional status and cognitive outcomes, controlling for covariates like sex, family care index, parental education, and income per needs, which may confound the relation between growth and neurocognitive development (Jensen, Berens & Nelson, 2017). Given the well-documented global impact of undernutrition on neurocognitive development, we hypothesized that growth faltering would be associated with distinct patterns in whole-brain EEG PSD and FC, and would predict poorer cognitive outcomes, with EEG PSD and/or FC mediating the relationship between nutritional status and cognitive performance.

## 2 METHODS

### 2.1 Participants

The study was conducted in Mirpur Ward 2,3, and 5, a densely populated urban slum in Dhaka, Bangladesh, characterized by challenging conditions such as limited literacy, unemployment, low income, and elevated malnutrition rates.

In 2022, 234 Bangladeshi infants (94% Bengali, 6% Bihari), aged 12 ± 1 months, were enrolled. Of these, 159 infants were diagnosed with moderate acute malnutrition (MAM), defined by a weight-for-length/height z-score (WHZ) between 2 and 3 standard deviations below the World Health Organization (WHO) reference mean, while 75 infants were well-nourished, with a WHZ greater than one standard deviation below the mean. Inclusion criteria excluded infants with chronic medical conditions or known congenital anomalies. This analysis is part of a randomized controlled trial on nutritional supplementation (see Shama et al., 2024 for details), but focuses on pre-intervention baseline measures at 12 months of age, including growth, EEG and cognition, treating all participants as a single cohort.

EEG data were obtained from 217 participants, with 140 (80 male, 60 female) meeting the criterion of at least 30 segments (1 minute) of clean resting-state EEG data post-artifact rejection. This final sample had an average household income of 4,528 taka per member per month (approximately $48.82, based on the 2022 conversion rate) with a standard deviation of 3,337 taka ($35.97). This income is below the World Bank’s international extreme poverty line of $2.15 per person per day, equivalent to $65.58 per month (Jolliffe et al., 2022). The final sample was representative of the original cohort in terms of ethnicity, sex, socioeconomic status, and growth measures.

Ethical approval was granted by the Research Review Committee (RRC) and the Ethical Review Committee (ERC) of the International Centre for Diarrheal Disease Research, Bangladesh (icddr,b) (protocol no: PR-21084), and the Institutional Review Board (IRB) of Boston Children’s Hospital, USA. Informed written consent was secured from the parents of all participating infants.

### 2.2 Measures

#### 2.2.1 Growth measures

Growth was assessed using measurements of supine length, weight, head circumference (HC), and mid-upper arm circumference (MUAC). Z-scores for length/height-for-age (HAZ), weight-for-age (WAZ), weight-for-length/height (WHZ), MUAC-for-age (MUACAZ), and HC-for-age (HCAZ) were calculated according to WHO standards.

These z-scores are standard indicators of undernutrition, categorized into stunting (HAZ < -2), underweight (WAZ < -2), and wasting (WHZ < -2). A MUAC measurement of less than 12.5 cm also indicates wasting. Moderate acute malnutrition (MAM) is a synonym for moderate wasting (defined by WHZ < -2 and ≥ -3 and/or MUAC between 11.5 cm and 12.5 cm). In our final sample (N = 140), 45 infants were stunted, 81 were underweight, and 89 were wasted/MAM.

For analysis, growth measures were treated as continuous variables to better represent nutritional health/status and the extent of linear growth faltering (see Perumal, Bassani & Roth, 2018). The mean z-scores for the final sample were: HAZ = -1.63 (SD = 0.97), WAZ = -1.9 (SD = 1.06), WHZ = -1.47 (SD = 1.03), MUACAZ = -1.28 (SD = 0.96), and HCAZ = -1.63 (SD = 0.91). Although MUAC is often used as an absolute measure to define acute malnutrition, we z-standardized MUAC to align with other growth indicators. This approach is supported by evidence suggesting that MUAC-for-age may be more effective than absolute MUAC in nutritional surveillance (Custodio et al., 2018).

#### 2.2.2 Cognitive assessment

Cognitive outcomes were evaluated using the Bayley Scales of Infant and Toddler Development, Fourth Edition (Bayley-IV), which includes subscales for cognitive, language (receptive and expressive), and motor (fine and gross) development. These assessments were administered by trained local research assistants and psychologists. The assessment items were translated and culturally adapted to increase relevance for the Bangladeshi context. Specifically, images in the item and action naming series of the language subscales were modified to reflect local cultural norms, such as depicting children in traditional Bangladeshi attire and substituting animals with local breeds. Objects and toys in the images were also adjusted, such as replacing a teddy bear with a pillow to better represent bedtime in the local context.

As the Bayley-IV is not standardized for the Bangladeshi population, U.S. norms were applied, limiting direct comparisons with Western populations. The scaled scores, adjusted for exact age at assessment using U.S. norms, were subsequently z-standardized within the sample. A composite/total score reflecting global cognitive development was calculated by averaging the z-scores across the five subscales.

#### 2.2.3 EEG data collection and processing

EEG data were collected using NetStation 4.5.4 and 128-channel HydroCel Geodesic Sensor Nets (HCGSN) with a NetAmps 300 amplifier (Electrical Geodesics, Inc., Eugene, OR, USA). Sessions were conducted in a dimly lit room, with participants seated on their parent’s lap and separated from the research staff by a curtain. A second staff member was present to assist with maintaining participant engagement and net positioning. The room lacked acoustic and electric shielding. Six paradigms were administered in each session, but this analysis focuses exclusively on the first: a quasi-resting-state session where participants watched a 3-minute screensaver video featuring toys, with accompanying sounds from the toys during the first half.

The EEG recordings were referenced online to a single vertex electrode, with data sampled at 500 Hz and impedances kept under 100 KΩ when possible. Preprocessing was performed offline in MatLab (R2023a) using the Harvard Automated Processing Pipeline for Electroencephalography (HAPPE) Version 4 (Gabard-Durnam et al., 2018). Steps included removing line noise with a 50 Hz updated CleanLine filter (Mullen, 2012), downsampling to 250 Hz, and applying a bandpass filter (1-50 Hz) with a Hamming windowed sinc FIR filter to minimize muscle and movement artifacts. Channels prone to artifacts were identified and rejected using joint probability criteria, and wavelet thresholding was applied for further artifact removal. Data were segmented into 2-second epochs, with segments exceeding amplitudes of |150 µV| or failing segment similarity criteria being discarded. Bad channels were interpolated, and data were re-referenced to the average of all channels. Inclusion in further analyses required at least 30 clean epochs (1/3 of the total session, i.e., 1 minute). The final sample had an average of 56.93 clean epochs (SD = 11.44).

EEG power decomposition was also performed using HAPPE 4. The Thomson Multitaper Method was employed for PSD estimation, applying a Fast Fourier Transform with multitaper windowing (Slepian tapers) to compute power (in µV²/Hz) for each 2-second segment and each channel. Total power (in µV²) was calculated via numerical integration across all frequencies within each of four age-appropriate frequency bands: theta (3-6 Hz), alpha (6-10 Hz), beta (10-22), and gamma (22-45). Whole-brain power for each frequency band for each participant was derived by averaging the total power in each band across all segments and channels. Although referred to as “PSD by frequency band”, this final measure technically represents total power, not power density. A natural logarithm transformation was applied to the power values to stabilize variance and normalize the distribution.

EEG FC analysis was conducted in the source space based on the procedures outlined by Xie et al. (2019a). Cortical sources were reconstructed using the Fieldtrip toolbox (Oostenveld et al., 2011), and age-matched MRI templates from the Neurodevelopmental MRI Database (Richards et al., 2016). Templates were segmented into component materials, and a Finite Element Method (FEM) model was constructed with gray matter as source volumes at a 6 mm grid resolution. The lead field and spatial filter matrices were estimated based on the FEM model. Distributed source reconstruction of EEG time-series was conducted using exact low-resolution electromagnetic tomography (eLORETA – Pascual-Marqui et al., 2011). Source volumes were segmented into 48 cortical regions of interest (ROIs) based on the LPBA40 brain atlas (Shattuck, et al., 2008), and source activities for each ROI were derived by averaging reconstructed time-series within the source volumes surrounding each ROI’s centroid. Subsequent FC analysis used the same frequency bands as PSD estimation: theta (3-6 Hz), alpha (6-10 Hz), beta (10-22), and gamma (22-45). Whole-brain FC between the 48 ROIs was estimated using the weighted phase lag index (wPLI), a measure of phase-to-phase synchrony that minimizes the noise influence by weighting phase differences based on the magnitude of leads and lags. To control for the varying segment numbers per participant after segment rejection, 30 segments were randomly selected in 50 iterations to calculate the average wPLI, producing 48x48 weighted adjacency matrices for each frequency band and each participant. Fisher’s z-transformation was applied to improve normality, and a sparsity threshold of 0.5 was employed to retain the strong but eliminate the weak or noise connections. The 48 ROIs were further grouped into the four lobes based on Shattuck et al. (2008) – frontal (F), temporal (T), parietal (P), and occipital (O) – and FCs within and between the four lobes (FF, FT, FP, FO, TT, TP, TO, PP, PO, OO) were calculated.

#### 2.2.4 Covariates

Sex assigned at birth, a family care index, maternal and paternal years of education, as well as income per needs were included as covariates in the final mediation models described below due to their potential association with children’s growth and their possible influence on EEG measures and cognitive outcomes. Given the sex imbalance in our sample and recent evidence indicating that undernutrition in children under five may disproportionately affect boys (Thurstans et al., 2020), including sex as a covariate was particularly important. Family caregiving activities and amount of stimulation at home were evaluated through interviews with the primary caregiver employing the Family Care Indicators (FCI) (Hamadani et al., 2010). The FCI encompasses five subscales: four assessing various aspects of home stimulation, such as the availability of play materials, books, magazines, and newspapers, and one “play activities” subscale capturing the frequency of stimulating interactions between the child and caregivers over the preceding three days. A sum-score from these subscales was calculated to form the family care index. Parental years of education and income per needs (defined as monthly household income divided by the number of household members) were included as indicators of childhood socioeconomic status (SES) (McLoyd, 1998).

### 2.3 Statistical analyses

All statistical analyses were performed using custom MatLab (R2023a) scripts and IBM SPSS Statistics (Version 29). Spearman correlation analyses were conducted to examine the relationships between growth measures (HAZ, WAZ, WHZ, MUACAZ, and HCAZ) – indicators of nutritional status – and whole-brain EEG PSD and FC across different frequency bands (theta, alpha, beta, and gamma). To control for type I error, correlations were adjusted for multiple comparisons using a 5% false discovery rate (FDR) threshold (Benjamini & Hochberg, 1995). Shapley value regression models, implemented in SPSS using the STATS RELIMP extension, were employed to determine the relative importance of each growth measure in explaining variance in EEG PSD and FC across the frequency bands (see Lipovetsky & Conklin, 2001). The relationships between EEG measures and cognitive outcomes (total Bayley scores), as well as between growth measures and cognitive outcomes, were also assessed using FDR-corrected Spearman correlations. Finally, mediation analyses were conducted using Hayes’ PROCESS macro (Model 4) in SPSS to explore potential indirect effects of nutritional status on cognitive outcomes via brain activity and FC measures (Hayes, 2017). These analyses focused on growth, and EEG measures that were significant in the initial Spearman correlations – both between growth and EEG measures, and between EEG measures and cognitive outcomes. When multiple growth measures were significant for a given EEG measure, the one with the highest Shapley value was selected. The mediation models evaluated the indirect effect of the selected growth measure on total Bayley scores through the selected EEG measure, controlling for covariates such as sex, family care index, maternal and paternal education, and income per needs. The significance of indirect effects was tested using bootstrapping with 5,000 samples, with 95% percentile bootstrap confidence intervals used to determine the robustness of the mediation effects.

## 3 RESULTS

### 3.1 Undernutrition and the brain

#### 3.1.1 Child growth and EEG PSD

To investigate the association between growth measures (indicators of nutritional status) and whole-brain EEG PSD in different frequency bands, Spearman correlations were conducted, with p-values adjusted using a 5% FDR threshold (see Figure 1). All five growth measures (HAZ, WAZ, WHZ, MUACAZ, and HCAZ) showed positive correlations with gamma (22-45 Hz) PSD. Beta (10-22 Hz) PSD was significantly positively correlated with all growth measures except HCAZ. In contrast, theta (3-6 Hz) and alpha (6-10 Hz) PSD showed significant positive correlations with HAZ and WAZ only; however, the association between WAZ and theta PSD did not withstand FDR correction. Overall, higher growth measure z-scores, reflecting better nutritional health, were associated with higher whole-brain EEG PSD, although the strength of this relationship varied by frequency band and growth measure.

**Figure 1:**
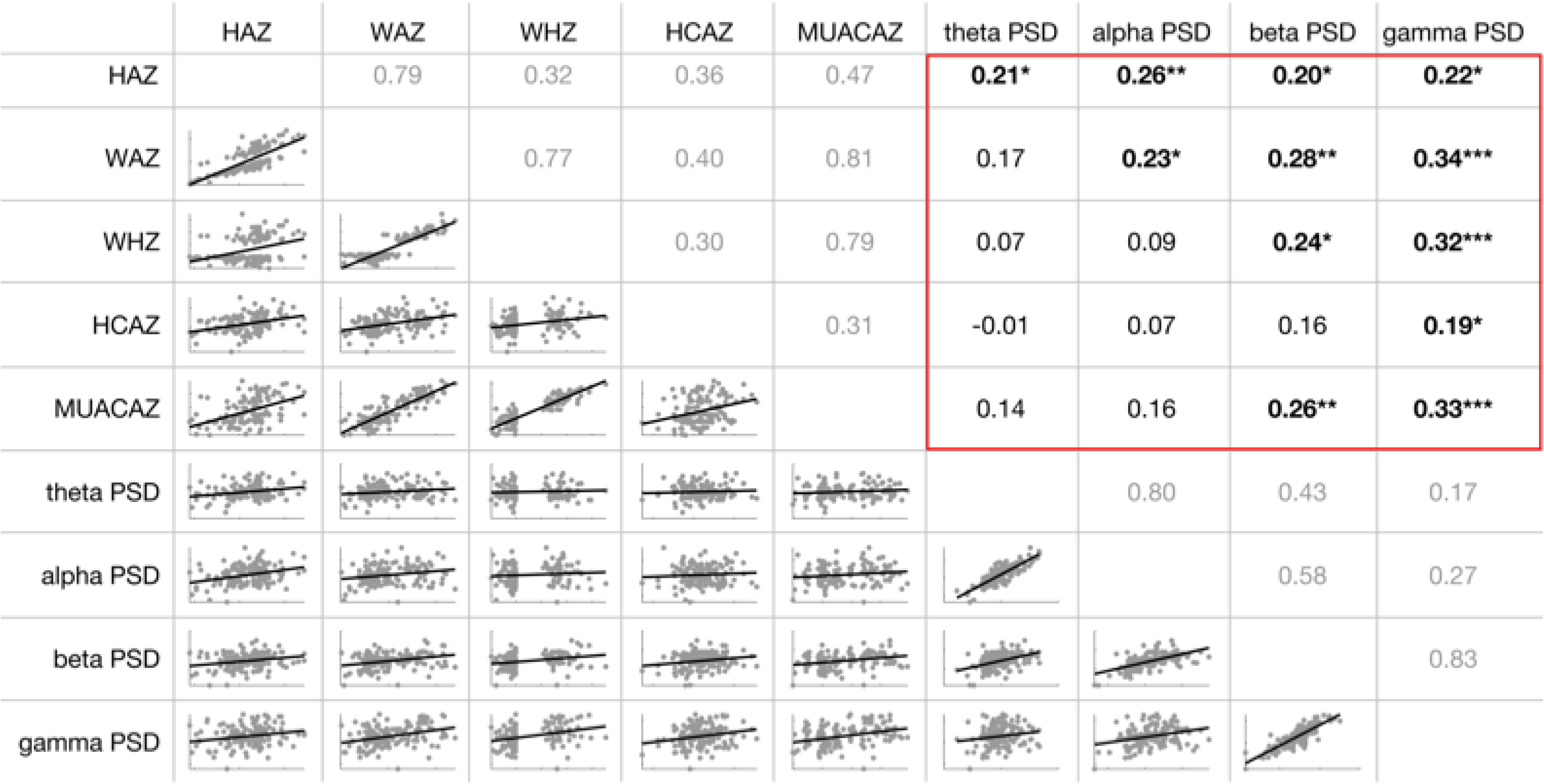
Correlation matrix illustrating the relationship between growth measures (HAZ, WAZ, WHZ, HCAZ, and MUACAZ) and whole-brain PSD in the frequency bands of interest (theta, alpha, beta, and gamma). The lower triangular matrix displays bivariate scatter plots with fitted linear regression lines for each pair. The upper triangular matrix presents Spearman correlation coefficients (ρ-values), with significance levels after FDR correction, indicated by symbols: ***q<0.001, **q<0.01, and *q<0.05. Correlations between any growth measure and any PSD measure are outlined in red, as these were the only correlations tested for significance and included in the FDR correction.

To assess the relative importance of different growth measures for EEG PSD across frequency bands, Shapley regression models were employed. Growth measures accounted for 13% of the variance in gamma PSD (R^2^=0.13). Shapley values (SVs – scaled to 100%) indicated that WAZ was the most influential growth measure for gamma PSD at 12 months (SV = 0.26), closely followed by MUACAZ and WHZ (both SV = 0.25), while HCAZ (0.15) and HAZ (SV = 0.09) appeared less influential. A similar pattern was observed for beta PSD (R^2^ = 0.11), where WAZ (SV = 0.28) and MUACAZ (SV = 0.23) explained the majority of the variance, with WHZ (SV = 0.19), HAZ (SV = 0.17), and HCAZ (SV = 0.14) contributing less. For alpha (R^2^ = 0.1) and theta (R^2^ = 0.08) PSD, HAZ emerged as the most important predictor (SV = 0.51 for both bands), markedly outweighing the contributions of WAZ (alpha: SV = 0.28, theta: SV = 0.26), WHZ (alpha: SV = 0.1, theta: SV = 0.13), MUACAZ (alpha: SV = 0.09, theta: SV = 0.09), and HCAZ (SV = 0.02 for both bands). These results show that growth measures explain more variance in EEG PSD at higher frequency bands, with HAZ being most predictive for theta and alpha PSD, and WAZ for beta and gamma PSD.

Figure 2 is visualizing whole-brain PSD across all analyzed frequency bands for the entire sample (Figure 2a), as well as separately for groups categorized by low and high HAZ (Figure 2b) and WAZ (Figure 2c) – the two growth measures identified as most relevant for PSD in the Shapley regression analysis. The categories for low and high HAZ and WAZ (< -2 vs. ≥ -2) were defined according to WHO criteria for undernutrition, where HAZ < -2 indicates stunting and WAZ < -2 indicates wasting. The figure shows differences in both the intercept and slope of whole-brain PSD between undernourished and well-nourished infants. These differences are observable across the entire power spectrum, with more pronounced effects at higher frequencies, particularly for low versus high WAZ.

**Figure 2:**
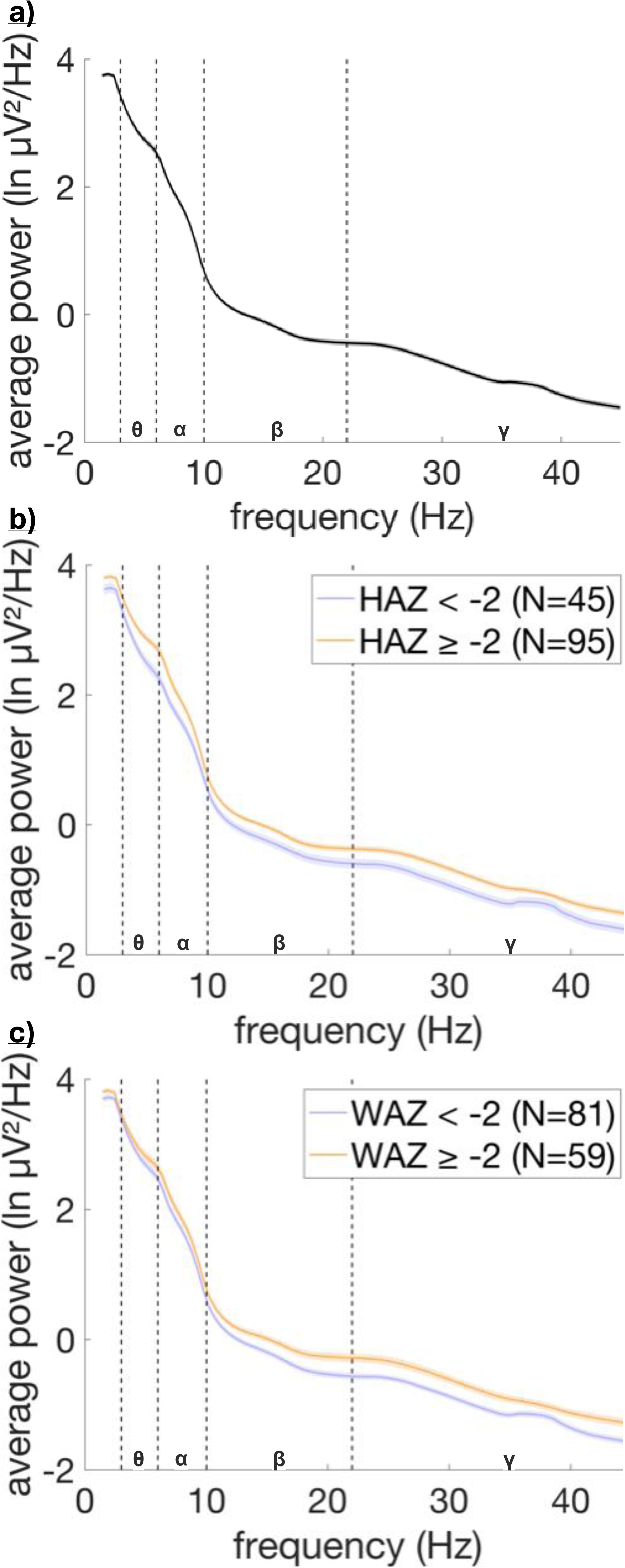
Average whole-brain power spectral density (a) across all 140 participants, (b) separately for participants with HAZ < -2 (N = 45) and HAZ ≥ -2 (N = 95), and (c) separately for participants with WAZ < -2 (N = 81) and WAZ ≥ -2 (N = 59). The shaded areas represent the standard errors of the mean for all participants in (a) and within each group in (b) and (c).

#### 3.1.2 Child growth and EEG FC

FDR-corrected Spearman correlations between growth measures and whole-brain EEG FC in different frequency bands (see Figure 3) initially indicated a significant negative association between MUACAZ and gamma FC, suggesting that infants with lower MUACAZ exhibit stronger whole-brain FC. However, this did not withstand FDR correction. Ultimately, no significant associations were found between any growth measures and whole-brain FC in any frequency band. To further explore the potential relationship between growth and gamma FC, we analyzed FC within and between the four lobes (i.e., FF, FT, FP, FO, TT, TP, TO, PP, PO, OO) in the gamma band. This analysis also revealed no significant relationships after FDR correction (see Supplementary Figure 1).

**Figure 3:**
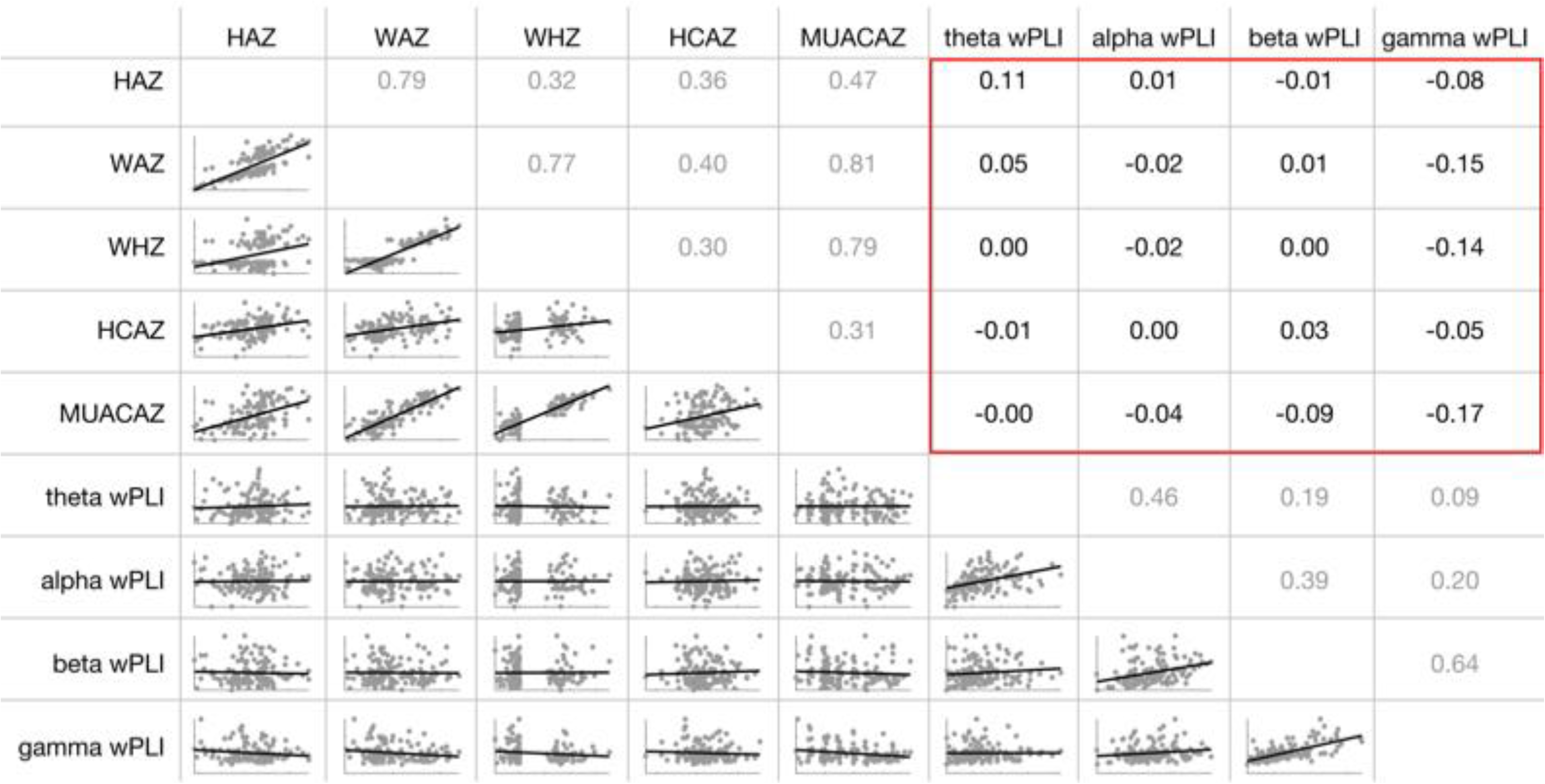
Correlation matrix illustrating the relationship between growth measures (HAZ, WAZ, WHZ, HCAZ, and MUACAZ) and whole-brain FC (estimated via wPLI) in the frequency bands of interest (theta, alpha, beta, and gamma). The lower triangular matrix displays bivariate scatter plots with fitted linear regression lines for each pair. The upper triangular matrix presents Spearman correlation coefficients (ρ-values), with significance levels after FDR correction, indicated by symbols: ***q<0.001, **q<0.01, and *q<0.05. Correlations between any growth measure and any FC measure are outlined in red, as these were the only correlations tested for significance and included in the FDR correction.

A Shapley regression was performed to evaluate the relative importance of various growth measures for whole-brain gamma FC, which showed a trending correlation with growth. Growth measures explained 5% of the variance in gamma FC (R^2^=0.05). Shapley values indicated that MUACAZ was the most influential predictor of gamma FC at 12 months (SV = 0.44), markedly exceeding WAZ (SV = 0.22), WHZ (SV = 0.18), HAZ (SV = 14), and HCAZ (SV = 0.02), which had minimal influence.

Whole-brain FC “spectral density” across all frequency bands is displayed in Figure 4, both for the entire sample (Figure 4a) and separately for groups categorized by low and high MUACAZ (Figure 4b). MUACAZ was selected for this comparison as it was identified as the most relevant growth measure for gamma FC in the Shapley regression analysis – the only frequency band where FC potentially correlated with growth. Low and high MUACAZ groups were defined as < -2 and ≥ -2, consistent with the categorization used for HAZ and WAZ above. The figure aligns with the correlation matrix, showing no discernible differences in whole-brain FC between undernourished and well-nourished infants in the lower frequency bands (delta, theta, and alpha). The trajectories are nearly identical within these bands, with overlapping peaks around 2 Hz, 4.5 Hz and 7.5 Hz. In contrast, the higher frequency bands, particularly beta and gamma, show descriptively higher FC levels in the malnourished group. Despite this, the primary peak around 37 Hz is evident in both groups, with no noticeable difference.

**Figure 4:**
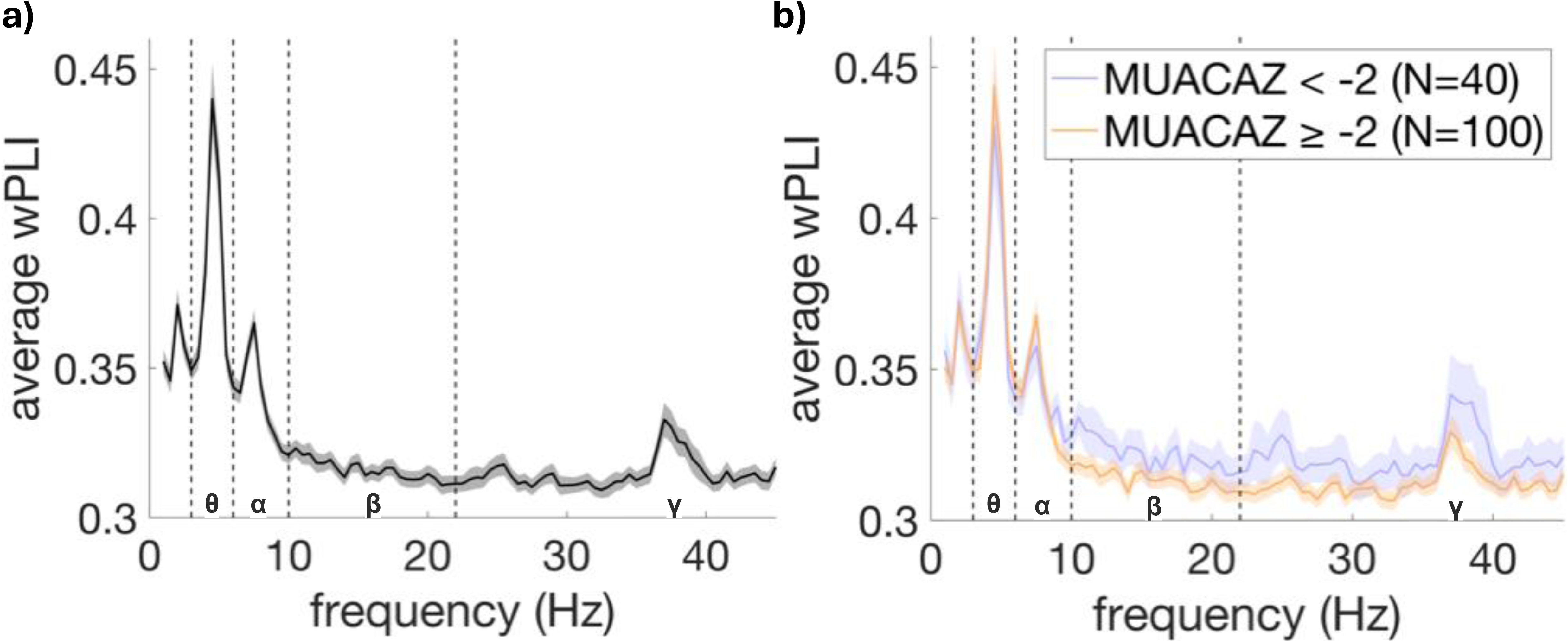
Average whole-brain functional connectivity (estimated via wPLI) “spectral density” (a) across all 140 participants, and (b) separately for participants with MUACAZ < -2 (N = 40) and MUACAZ ≥ -2 (N = 100). The shaded areas represent the standard errors of the mean for all participants in (a) and within each group in (b).

### 3.2 Brain and cognition

#### 3.2.1 EEG PSD and Bayley IV

To examine the relationship between whole-brain PSD in different frequency bands and cognitive outcomes, specifically the total Bayley scores, FDR-corrected Spearman correlations were conducted (see Figure 5). Initial analyses indicated significant positive correlations between PSD in all frequency bands except gamma and Bayley scores, suggesting that infants with higher PSD amplitudes tend to have higher cognitive scores. However, after FDR correction, only the correlations with theta and alpha PSD remained significant. This suggests that, contrary to growth measures, cognitive outcomes exhibit stronger correlations with PSD in lower frequency bands.

**Figure 5:**
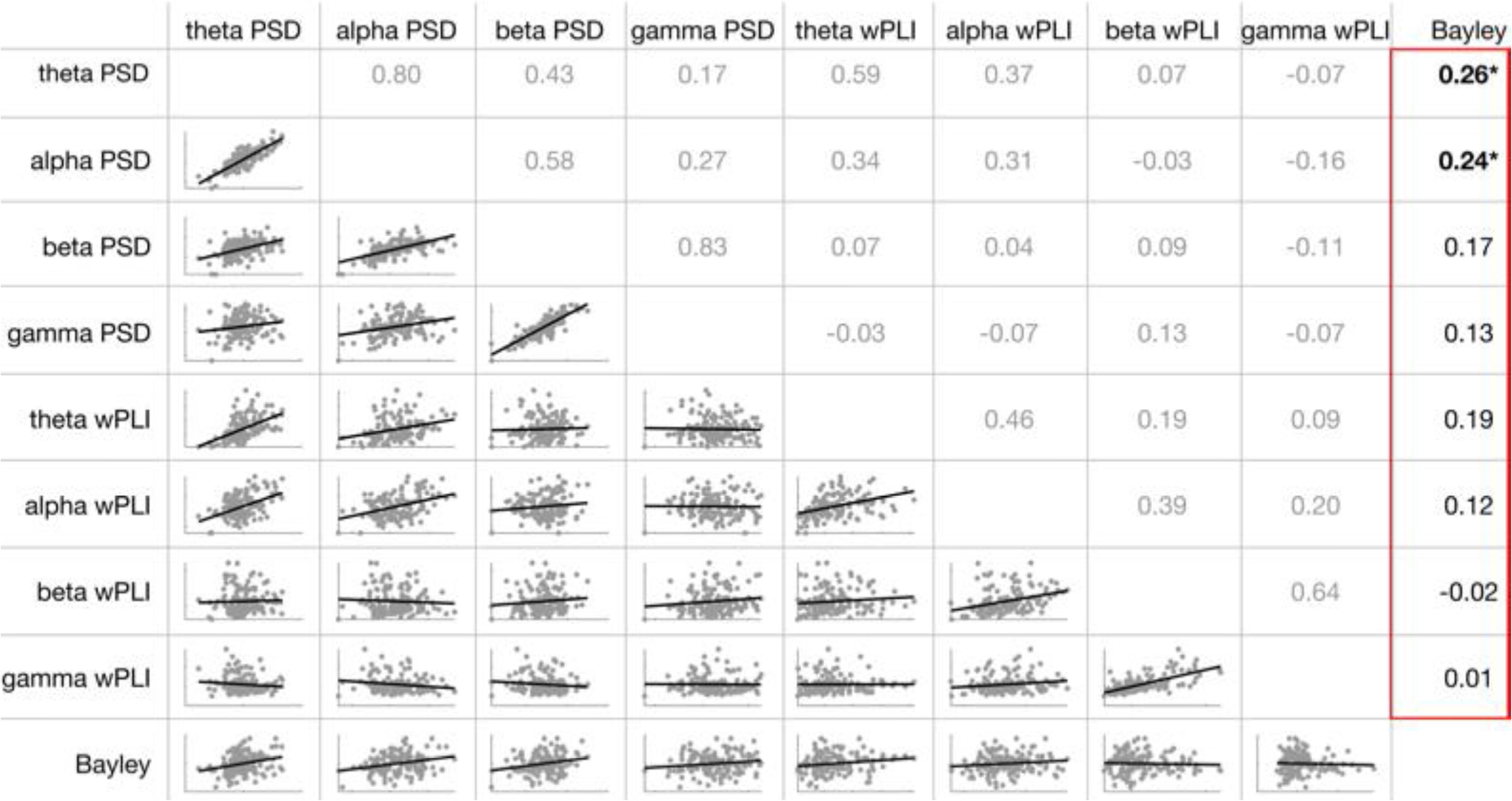
Correlation matrix illustrating the relationship between whole-brain PSD as well as FC (estimated via wPLI) in the frequency bands of interest (theta, alpha, beta, and gamma) and total Bayley scores. The lower triangular matrix displays bivariate scatter plots with fitted linear regression lines for each pair. The upper triangular matrix presents Spearman correlation coefficients (ρ-values), with significance levels after FDR correction, indicated by symbols: ***q<0.001, **q<0.01, and *q<0.05. Correlations between any EEG measure and Bayley scores are outlined in red, as these were the only correlations tested for significance and included in the FDR correction.

#### 3.2.2 EEG FC and Bayley IV

The FDR-corrected Spearman correlation models used for PSD were also applied to assess the relationship between FC and total Bayley scores (see Figure 5). Initial analyses indicated a significant positive association between theta FC and Bayley scores, suggesting that stronger FC might be linked to higher cognitive scores. However, this association did not remain significant after FDR correction. Ultimately, no significant correlations were found between FC in any frequency band and Bayley scores.

### 3.3 Mediation model linking growth to cognition via EEG PSD

Figure 6 illustrates the FDR-corrected Spearman correlations between growth measures (HAZ, WAZ, WHZ, HCAZ, and MUACAZ) and total Bayley scores. The analysis indicated that none of the growth measures were significantly associated with Bayley scores.

**Figure 6:**
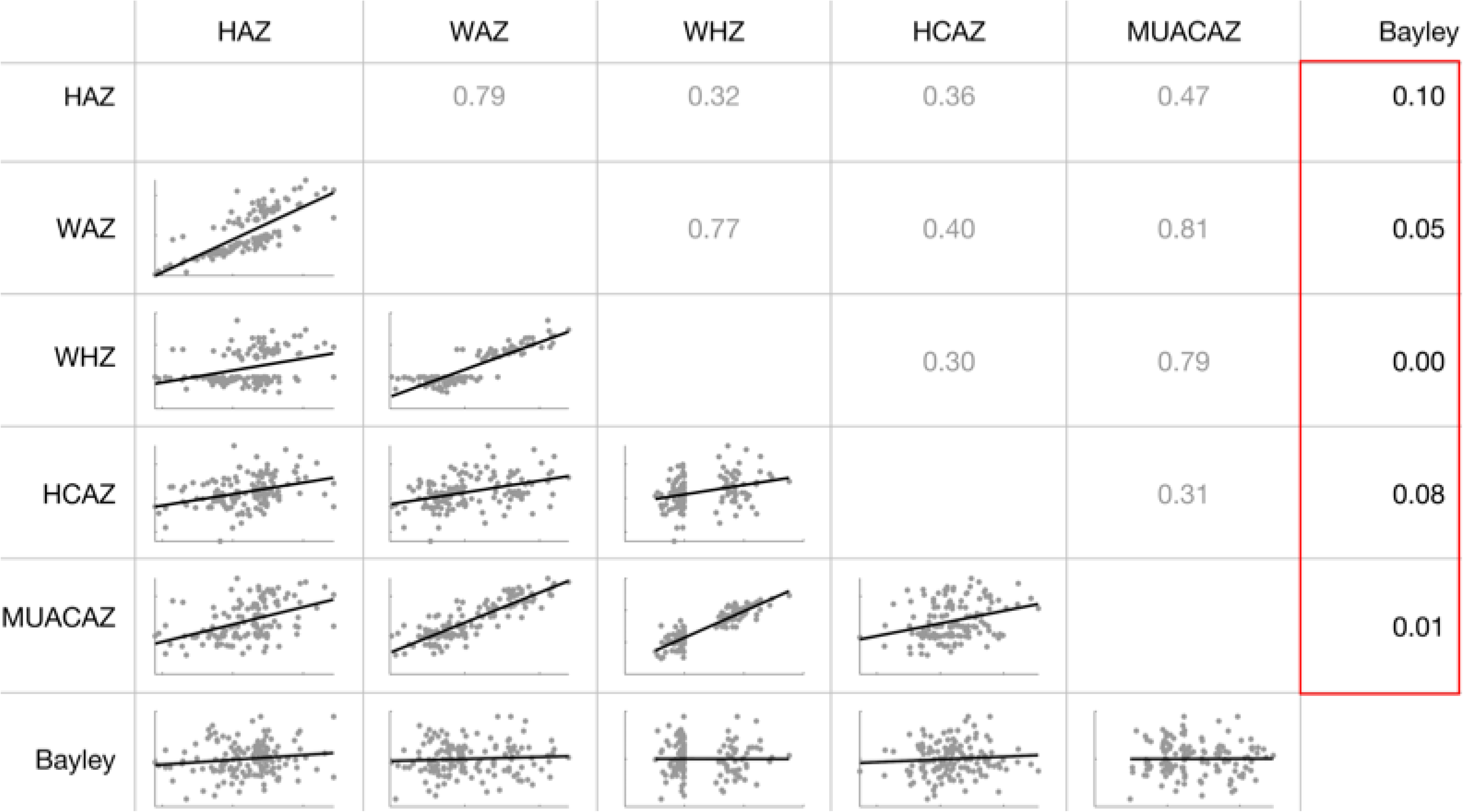
Correlation matrix illustrating the relationship between growth measures (HAZ, WAZ, WHZ, HCAZ, and MUACAZ) and total Bayley scores. The lower triangular matrix displays bivariate scatter plots with fitted linear regression lines for each pair. The upper triangular matrix presents Spearman correlation coefficients (ρ-values), with significance levels after FDR correction, indicated by symbols: ***q<0.001, **q<0.01, and *q<0.05. Correlations between any growth measure and Bayley scores are outlined in red, as these were the only correlations tested for significance and included in the FDR correction.

Given the significant correlations between growth measures (particularly HAZ in this case) and EEG PSD, as well as between EEG PSD and total Bayley scores – especially in the theta and alpha bands – we conducted a mediation analysis to explore a potential indirect effect of growth (as an indicator of nutritional status) on cognitive outcomes (see Figure 7). Using Hayes’ PROCESS Model 4, we evaluated the indirect effect of HAZ on total Bayley scores through theta and alpha EEG PSD, controlling for covariates such as sex, family care index, maternal and paternal education, and income per needs.

**Figure 7:**
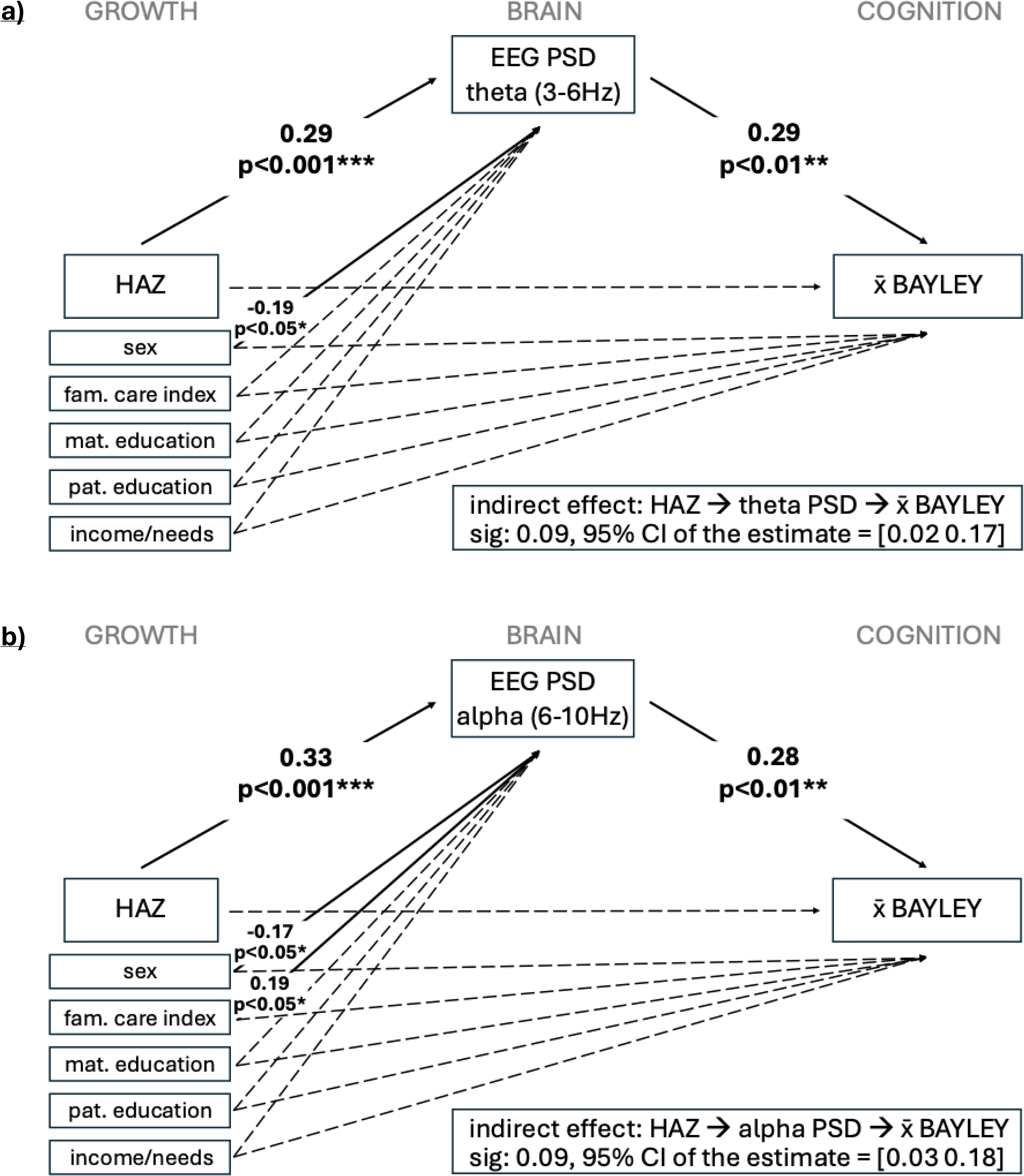
Mediation models of HAZ on total Bayley scores through **(a)** theta PSD and **(b)** alpha PSD, controlling for sex, a family care index, maternal and paternal education, as well as the income per needs ratio. The diagrams illustrate the mediation models tested using Hayes’ PROCESS Model 4. Standardized path coefficients (β) are displayed along the arrows for significant paths, along with their respective p-values (***p<0.001, **p<0.01, *p<0.05), indicating the strength and direction of the relationships. The coefficient and 95% percentile confidence interval for the indirect effect, based on a bootstrap analysis with 5,000 samples, are presented in the black box at the bottom of each diagram.

HAZ had a significant positive effect on both theta (Figure 7a) and alpha PSD (Figure 7b), suggesting that better nutritional health, indicated by higher HAZ, is associated with increased theta and alpha PSD. Additionally, sex had a significant effect on both theta and alpha PSD, with male infants exhibiting higher PSD amplitudes. The family care index also significantly affected alpha PSD, with higher levels of family care linked to increased PSD. Further, both theta and alpha PSD had significant positive effects on the Bayley score, suggesting that higher PSD in these frequency bands is associated with better cognitive outcomes. No direct effect of HAZ on the Bayley score was observed. Bootstrap analysis with 5,000 samples confirmed the significance of the indirect effects in both models, with 95% confidence intervals excluding zero, indicating a robust mediation effect. Thus, HAZ exerts an effect on the Bayley score indirectly through theta as well as alpha PSD.

## 4 DISCUSSION

### 4.1 Theoretical mechanisms

This study provides evidence that various growth measures, used to assess nutritional status, are positively associated with variations in brain activity (PSD) across frequency bands as early as 12 months of age. There is also a preliminary indication of an association between better nutritional status and reduced brain connectivity (FC) in the gamma band, though these findings did not withstand FDR correction. Both PSD and FC have been widely used to study neural maturation in typically developing children (e.g., Gasser et al., 1988; Bathelt et al., 2013) and those exposed to early adversity (e.g., Valdés-Sosa et al., 2018; Xie et al., 2019a). PSD measures brain activity, while FC provides insights into the organization and functionality of brain networks. The observed correlations between growth and whole-brain PSD and FC may reflect disrupted brain function, aligning with established literature on neural development (Georgieff et al., 2018).

The development of the power spectrum over time is well-documented. Earlier studies (Gasser et al., 1988) have shown a decrease in absolute power across all frequency bands except alpha during childhood (6-17 years), with relative power shifting from slower to faster bands. More recently, research has focused on two components of the power spectrum: the periodic (oscillatory) component, which reflects rhythmic brain activity, and the aperiodic component, which represents the non-oscillatory neuronal spiking activity. In line with previous findings, newer studies indicate that the aperiodic slope of the power spectrum decreases with age, reflecting changes in the excitation/inhibition (E/I) balance in children aged 4 to 12 years (Hill et al., 2022). This decline continues into adulthood, where flatter slopes are linked to neural noise and age-related cognitive decline (Voytek et al., 2015). Additionally, flattened aperiodic slopes are often observed alongside decreased aperiodic offsets with age, likely reflecting reductions in cortical grey matter volume during childhood, driven by maturational processes like synaptic pruning (see Couperus & Nelson, 2006). Contrasting these patterns observed in children and adults, a recent study (Wilkinson et al., 2024) demonstrated significant increases in both aperiodic slope and offset during infancy, especially within the first year. The early increase in offset likely reflects rapid brain volume expansion and synaptogenesis known to occur during this period. The increase in slope during infancy may indicate a reduction in the E/I ratio, aligning with the prolonged maturation of inhibitory networks in humans (Paredes et al., 2016). While our analyses did not explicitly examine periodic and aperiodic components, the observed correlations with growth across the whole power spectrum – along with more pronounced differences between well-nourished and malnourished infants at higher frequencies – suggest that malnourished infants exhibit lower offsets but steeper slopes (Figures 1 and 2). The typical age-related increases in offset and slope until around 16 months (Wilkinson et al., 2024) could imply that lower offsets in malnourished infants represent delayed brain development, particularly in terms of brain volume expansion and synaptogenesis. Conversely, steeper slopes seen in malnourished infants could be misleading if interpreted as indicative of more mature development. However, it’s crucial to note that the developmental changes in the aperiodic slope are gradual, with less pronounced daily shifts within the first year of life compared to aperiodic offset. Additionally, steeper slopes usually indicate reduced E/I balance, suggesting a deficit in excitatory activity. E/I imbalances can signal disruptions in typical brain development. Furthermore, periodic power changes across frequency bands during infancy also differ from later stages. Wilkinson et al. (2024) found that during the first year of life, periodic power increased across most frequency bands except theta and gamma, which already peaked around 6 months. In contrast, malnourished infants showed reduced periodic power, potentially reflecting decreased neuronal firing or synchronization, which are indicative of delayed or atypical neural development. High-frequency activity is energetically demanding, and reduced power in these bands may be directly linked to compromised energy metabolism due to malnutrition.

The developmental trajectory of EEG FC during infancy remains underexplored, making it difficult to fully understand normative patterns. While fMRI studies (e.g., Gao et al., 2015) provide insights into brain connectivity, longitudinal studies are needed to trace typical neural network development (Cosío-Guirado, 2024). Like PSD, FC generally increases in early development, reflecting synaptogenesis and brain volume expansion. On the other hand, declines in FC signify processes like synaptic pruning and axonal myelination, which promote network specialization and neural efficiency. These processes occur heterochronously in different cortical regions during the first year, reflecting rapid network refinement. As local connections weaken, long-distance connections are strengthened and myelinated, increasing integration and improving neural communication efficiency across the brain (Ma & Zhang, 2021). Data from Xie et al. (2019a) on FC “spectral density” of children aged 6 and 36 months suggest a decrease in whole-brain FC across all frequency bands except alpha, mirroring later PSD developmental trajectories (see Gasser et al., 1988). With this preliminary evidence of a normative decline in FC during this period, the elevated gamma FC observed in malnourished infants may be suggestive of delayed brain maturation, particularly in myelination and the early stages of synaptic pruning. This could imply that local connections have not sufficiently weakened, or distal connections have not yet reached age-appropriate myelination, resulting i2n increased whole-brain FC.

Furthermore, our study provides evidence that brain activity, as measured by PSD, particularly in lower frequency bands (theta and alpha, with preliminary evidence for beta which did not survive FDR correction), is positively correlated with cognitive outcomes, as measured by the total Bayley score. Additionally, there is preliminary evidence suggesting that FC in the theta band may also be positively related to cognitive outcomes at this age, although these findings did not withstand FDR correction. These results align with the known role of theta and alpha oscillations in cognitive functions like memory and attention (Klimesch, 1999; Orekhova et al., 2001, 2006), indicating that atypical PSD (and FC) patterns in these frequency bands may contribute to deficits in key cognitive domains.

This adds to the growing literature linking atypical PSD or FC patterns due to biological adversity to cognitive deficits (e.g., Gabard-Durnam et al., 2019; Orekhova et al., 2014; Xie et al., 2019a). Interestingly, both PSD and preliminary FC correlations with cognitive outcomes were positive, with the FC finding contrasting with previous results for older children (Xie et al., 2019a). Lower PSD might be linked to poorer cognitive outcomes due to decreased neuronal firing, reduced synchronization, or E/I imbalance. Similarly, poorer cognitive outcomes linked to decreased FC in malnourished infants may result from underdeveloped integration of large-scale networks. The associations between brain activity, as well as connectivity, and cognitive outcomes were more pronounced in the lower frequency bands for PSD and were only observed in the lower frequencies (theta) for FC. This finding is noteworthy because existing literature often links higher frequencies to cognitive outcomes (e.g., Benasich et al., 2008), and the effects of malnutrition were more pronounced in the higher frequencies for PSD and were only present in the higher frequencies (gamma) for FC at the same age. However, it is crucial to consider that the study by Benasich et al. (2008) found gamma power to be significantly related only to expressive language scores at 16 months, with broader cognitive relationships emerging only at 24 months, suggesting that the relevance of gamma oscillations for cognitive outcomes may increase later in development. Additionally, gamma activity is particularly susceptible to contamination by muscle and movement artifacts (Whitham et al., 2007), which could obscure relationships in the data. The dissociation, with nutritional status having a more pronounced impact on higher frequencies, while lower frequencies are more relevant for cognitive outcomes at 12 months, might explain the absence of a total effect of malnutrition on cognitive outcomes at this age.

While we did not find a total or direct effect of growth on cognitive outcomes at 12 months, a particularly compelling result is that HAZ indirectly affects Bayley scores through its effects on theta and alpha PSD. This suggests that cognitive impairments may primarily occur when malnutrition impacts brain function, which in turn affects cognitive outcomes. These results underscore the importance of neural activity as a critical pathway through which early adversity, including malnutrition, affects cognitive development. Importantly, our study highlights the potential of EEG as a sensitive tool for detecting PSD differences related to early adversity and predicting later cognitive outcomes, particularly in low-resource settings.

Additionally, sex and family care index were both associated with PSD, with male infants exhibiting higher PSD amplitudes and greater family care correlating with increased PSD. Given the multitude of factors interacting with child growth, researchers should recognize that brain and cognitive development deficits among infants in low- and middle-income countries are likely influenced by a complex interplay of factors (Nelson, 2017).

While this study provides robust evidence for associations between growth faltering, brain activity, and cognitive outcomes at 12 months, the corresponding evidence for FC is less conclusive. It is essential to recognize that the same experience can affect the brain differently depending on the developmental stage (Kolb and Gibb, 2011). One reason for the stronger associations observed with PSD compared to FC at this age might be that neural oscillatory activity, as captured by PSD, is more directly impacted by malnutrition. PSD, a potentially more immediate and sensitive marker, reflects energetically demanding brain activities that could be compromised in malnourished infants due to impaired energy metabolism. In contrast, FC, revealing the organization and functional dynamics of brain networks, may be a less sensitive measure for the early effects of malnutrition at 12 months, when these networks are still relatively immature. However, FC may become more informative at later developmental stages when these networks have further matured, as shown in studies of older children (Xie et al., 2019a). This is consistent with fMRI findings that while primary networks in neonates are topologically adult-like, higher-order networks remain incomplete and isolated but undergo consistent synchronization during the first two years of life (Gao et al., 2015).

This study used various growth measures to capture different facets of malnutrition and its potential impact on brain development and cognitive outcomes. Our findings highlight distinct relationships between specific growth and EEG measures, reflecting the complexity of how different aspects of malnutrition affect the developing brain. For instance, HAZ showed the strongest association with PSD in lower frequency bands, suggesting that especially chronic undernutrition may impact these foundational neural processes (see WHO, 2024b). In contrast, WAZ was most relevant for PSD in higher frequencies, potentially reflecting the impact of both acute and chronic nutritional deficits (see WHO, 2024b). Additionally, MUACAZ showed the strongest association with gamma FC, suggesting a potential link between body composition and neural network development (see Eaton-Evans, 2013). The differential associations between each growth measure and brain development suggest they may reflect different aspects of undernutrition, each affecting neural and cognitive development in distinct ways, potentially depending on the child’s age and developmental stage. Understanding the exact nutrient-level mechanisms driving these associations is crucial and remains a key area for future research.

### 4.2 Limitations

This study has some limitations that should be acknowledged. First, while we relied on growth measures as indicators of undernutrition, they may not fully capture all direct effects of undernutrition on the brain. Consequently, our study might underestimate the detrimental impact of malnutrition, as these measures do not encompass all aspects of nutritional health. Furthermore, the growth metrics used may capture more than just nutritional health, as they are correlated with multiple co-occurring factors related to severe adversity, including SES, sanitation, inflammation, and parental stress, which may independently affect neural and cognitive development (Xie et al., 2019b).

Second, our intervention study design may have introduced bias. The disproportionate representation of participants with WHZ values either between -2 and -3 (MAM intervention group, N=89) or above -1 (control group, N=51), resulting in a gap in the WHZ distribution, could limit the generalizability of our findings and affect the interpretation of WHZ as a predictor of brain and cognitive outcomes. This skewed and discontinuous WHZ distribution might also account for the less robust associations between WHZ and our brain measures, potentially obscuring the true relationship between WHZ and neurodevelopmental outcomes.

Lastly, the study may have been underpowered, limiting our ability to detect significant effects, especially given the multiple factors included in our statistical models. This limitation might explain some non-significant associations, such as those between growth measures, FC, and cognitive outcomes after FDR correction. To address these issues, data sharing and collaboration between multiple research sites are crucial to more comprehensively examine the contributions of different adversity aspects to child brain and cognitive development in low- and middle-income countries.

## 5 CONCLUSION

This study provides valuable insights into the complex relationships between nutritional health, brain development, and cognitive outcomes in infancy. Our findings demonstrate that nutritional status, as indicated by various growth measures, is already associated with distinct patterns of neural activity and connectivity by 12 months of age. Specifically, low HAZ, indicative of chronic undernutrition, emerged as the strongest predictor of decreased PSD in lower frequency bands (theta and alpha), while low WAZ was most closely associated with reduced power in higher frequencies (beta and gamma). Preliminary associations were also observed between low MUACAZ and increased gamma FC, although these did not survive FDR correction. Declines in certain neural markers, particularly theta and alpha PSD, with preliminary evidence (not surviving FDR) for beta PSD, were, in turn, associated with poorer cognitive outcomes at 12 months. Interestingly, a preliminary positive association between theta FC and cognitive outcomes was found (not surviving FDR), despite theta FC not being correlated with nutritional status. This dissociation – where nutritional status predominantly affects higher frequencies, while lower frequencies are more strongly linked to cognitive outcomes at 12 months – may explain the absence of a total effect of malnutrition on cognitive outcomes at this age. Importantly, however, our study reveals that HAZ indirectly influences Bayley scores through its effect on theta and alpha PSD. These findings deepen our understanding of the neural mechanisms through which growth faltering impacts cognitive development and highlight the potential for targeted interventions to mitigate the adverse effects of malnutrition in children from low- and middle-income countries.

## Data Availability

All data produced in the present study are available upon reasonable request to the authors

## ACKNOWLEDGEMENTS

The authors wish to express their gratitude to the participants from Mirpur, Dhaka, Bangladesh, for their invaluable contributions to this study. We also extend our thanks to the study team within the International Centre for Diarrheal Disease Research, Bangladesh (icddr,b), for their dedicated work in participant recruitment, sample collection, and assessments.

This work is supported by funding from the Wellcome Leap 1kD Program. We also acknowledge our core donors, Government of Bangladesh, and Canada for providing unrestricted support and commitment to icddr,b’s research effort.

## SUPPLEMENTS

**Supplementary Figure 1:**
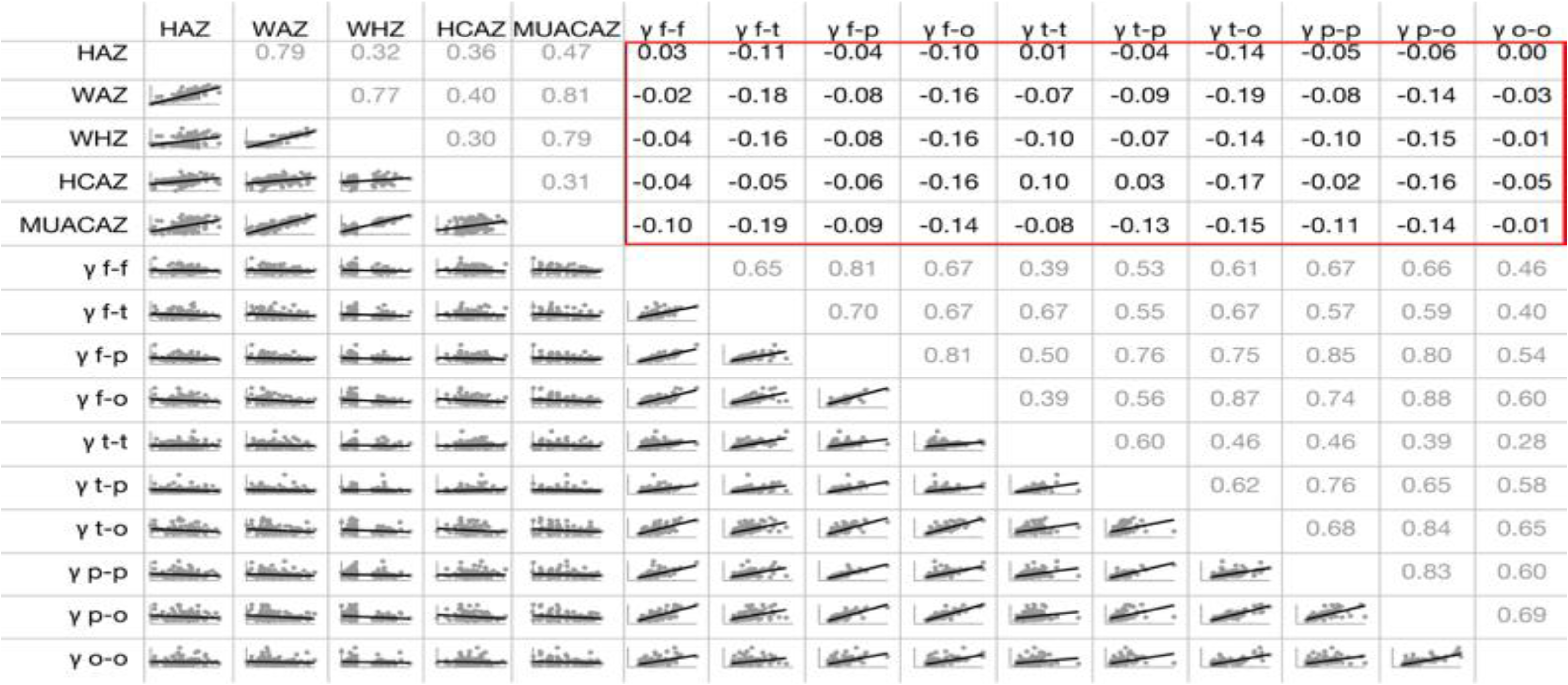
Correlation matrix illustrating the relationship between growth measures (HAZ, WAZ, WHZ, HCAZ, and MUACAZ) and gamma band (22-45 Hz) FC (estimated via wPLI) within and between the four lobes: frontal (F), temporal (T), parietal (P), and occipital (O), i.e., FF, FT, FP, FO, TT, TP, TO, PP, PO, OO. The lower triangular matrix displays bivariate scatter plots with fitted linear regression lines for each pair. The upper triangular matrix presents Spearman correlation coefficients ( ρ-values), with significance levels after FDR correction, indicated by symbols: ***q<0.001, **q<0.01, and *q<0.05. Correlations between any growth measure and any FC measure are outlined in red, as these were the only correlations tested for significance and included in the FDR correction.

